# ECG-Derived Synthetic Tissue Doppler Waveforms Differentiate Physiological Adaptations in Healthy Athletes from Pathological Patterns Associated with Mortality in Young Individuals

**DOI:** 10.1101/2025.08.18.25333913

**Authors:** Mehdi Kushkestani, Ankush D Jamthikar, Sabahat Bokhari, Jason P. Womack, Naveena Yanamala, Partho P Sengupta

**Author notes:** Shared contributions (Mehdi Kushkestani, Ankush D Jamthikar). **Corresponding Author** Partho P. Sengupta, MD, DM, FACC, FASE, Rutgers Robert Wood Johnson Medical School, Division of Cardiovascular Disease and Hypertension, 125 Patterson St, New Brunswick, NJ – 08901.

## Abstract

**Background:** Sudden cardiac death (SCD) in young individuals—especially athletes—remains difficult to diagnose due to overlapping physiological and pathological electrocardiogram (ECG)-related changes. Black-box artificial intelligence (AI) models can detect abnormal ECGs but often lack physiological interpretability. Tissue Doppler Imaging (TDI) provides functional insight into myocardial mechanics but requires echocardiographic imaging. This study investigates whether synthetic TDI features derived from standard ECGs can link electrical patterns to myocardial function, helping distinguish elite athletes, high-risk individuals with abnormal ECGs who died, and long-term survivors with normal ECGs.

**Methods:** Synthetic systolic, early, and late diastolic myocardial velocities (s′, e′, a′) and mitral annular displacement were generated from resting 12-lead ECGs using a previously validated Generative Adversarial Network (GAN) model. Data came from 28 Norwegian endurance athletes and a 233,770-person open-source community dataset, from which we identified high-risk (n = 21) and controls (n = 29) subjects. Logistic regression quantified the discriminative value of synthetic TDI parameters; model performance was assessed via area under the receiver operating characteristic curve (AUC).

**Results:** Athletes exhibited higher e′, e′/a′ ratios, and septal displacement than high-risk, consistent with enhanced diastolic compliance. High-risk showed reduced velocities and displacement, consistent with impaired relaxation and subclinical systolic dysfunction. Synthetic e′, e′/a′ ratio, and average maximum displacement differentiated athletes from high-risk with ORs of 1.51 (95% CI: 1.04 – 2.20), 5.58 (95% CI: 1.32 – 23.14) and 28.90 (95% CI: 4.24 – 197.18), respectively. A multivariate model showed an OR of 24.66 (CI: 3.23 – 188.25).

**Conclusion:** ECG-derived synthetic TDI offers a physiologically interpretable link between ECG patterns and myocardial function, distinguishing benign athletic remodeling from patterns associated with mortality risk. As an explainability tool, it may provide functional context for AI-based ECG interpretation, supporting scalable, imaging-free screening in athletic and general populations.

## Introduction

Artificial intelligence (AI) has shown promise in detecting subtle electrocardiographic (ECG) abnormalities that are predictive of adverse outcomes; however, most models function as “black boxes,” offering limited insight into the underlying physiological mechanism^1^. For athletic populations, where benign ECG changes often mimic pathological patterns, this lack of interpretability can erode clinician trust and hinder adoption ^2^. Explainable AI (XAI) seeks to bridge this gap by producing domain-relevant, physiologically meaningful outputs that clarify the basis for model predictions^1^.

Cardiovascular disease (CVD) remains the leading cause of death worldwide, accounting for nearly one-third of U.S. mortality and costing over $360 billion annually^3,4^. While less frequent, sudden cardiac death (SCD) in young adults—particularly athletes—remains devastating, with an incidence of 1 in 37,000 to 1 in 80,000 athletes annually, often without preceding symptoms^5–7^. Distinguishing mild athletic remodeling from early-stage, high-risk disease is a persistent challenge, as standard ECG findings such as T-wave inversion, ST-segment changes, and high QRS voltage overlap between physiological adaptation and electrical vulnerability^8,9^. This underscores the need for scalable, imaging-free tools that link ECG findings to functional cardiac performance.

Tissue Doppler Imaging (TDI) is a clinical gold standard for assessing myocardial mechanics, providing precise, motion-based measures of systolic and diastolic function^10–12^. However, echocardiographic acquisition requirements limit its widespread use. AI offers a potential solution by reconstructing functional parameters directly from ECG data. Our recently validated deep learning model synthesizes TDI features from standard 12-lead ECGs, leveraging electromechanical coupling to estimate myocardial velocities and displacement. In 585 patients, synthetic TDI parameters showed strong correlation with echocardiographic measurements, achieving AUCs of 0.80 and 0.81 for detecting diastolic and systolic dysfunction, respectively^1^.

TDI has been shown to distinguish between physiological remodeling and subclinical pathology. Elite athletes demonstrate higher E/A ratios and lower heart rates compared with healthy controls ^13^, whereas genotype-positive, phenotype-negative hypertrophic cardiomyopathy (HCM) patients display increased wall thickness and reduced E/A ratios ^14^. Nagueh et al. (2008) further confirmed TDI’s capacity to differentiate pathological from physiological hypertrophy in borderline cases ^15^. Extending this capability to synthetic, ECG-derived TDI provides a scalable and interpretable means of functional assessment, transforming an AI model’s prediction from a statistical label into a physiologically grounded explanation.

This pilot study evaluates whether ECG-derived synthetic TDI features can differentiate physiological adaptation in elite endurance athletes from abnormal ECG patterns associated with premature death in young individuals, as well as from healthy controls with normal ECGs. By embedding functional electromechanical signatures into ECG analysis, we aim to establish synthetic TDI not only as a diagnostic tool but also as an explainability framework that links ECG morphology to cardiac function, providing preliminary effect sizes for future validation studies.

## Methods

### Study Design and Population

A total of 77 participants were included in this pilot study (**Central Illustration**), divided into three phenotypic groups: elite endurance athletes (n = 27), individuals with abnormal ECGs who subsequently died during follow-up (n = 21), and controls with normal ECGs and no documented cardiac events during at least five years of follow-up (n = 29).

**Central Illustration:**
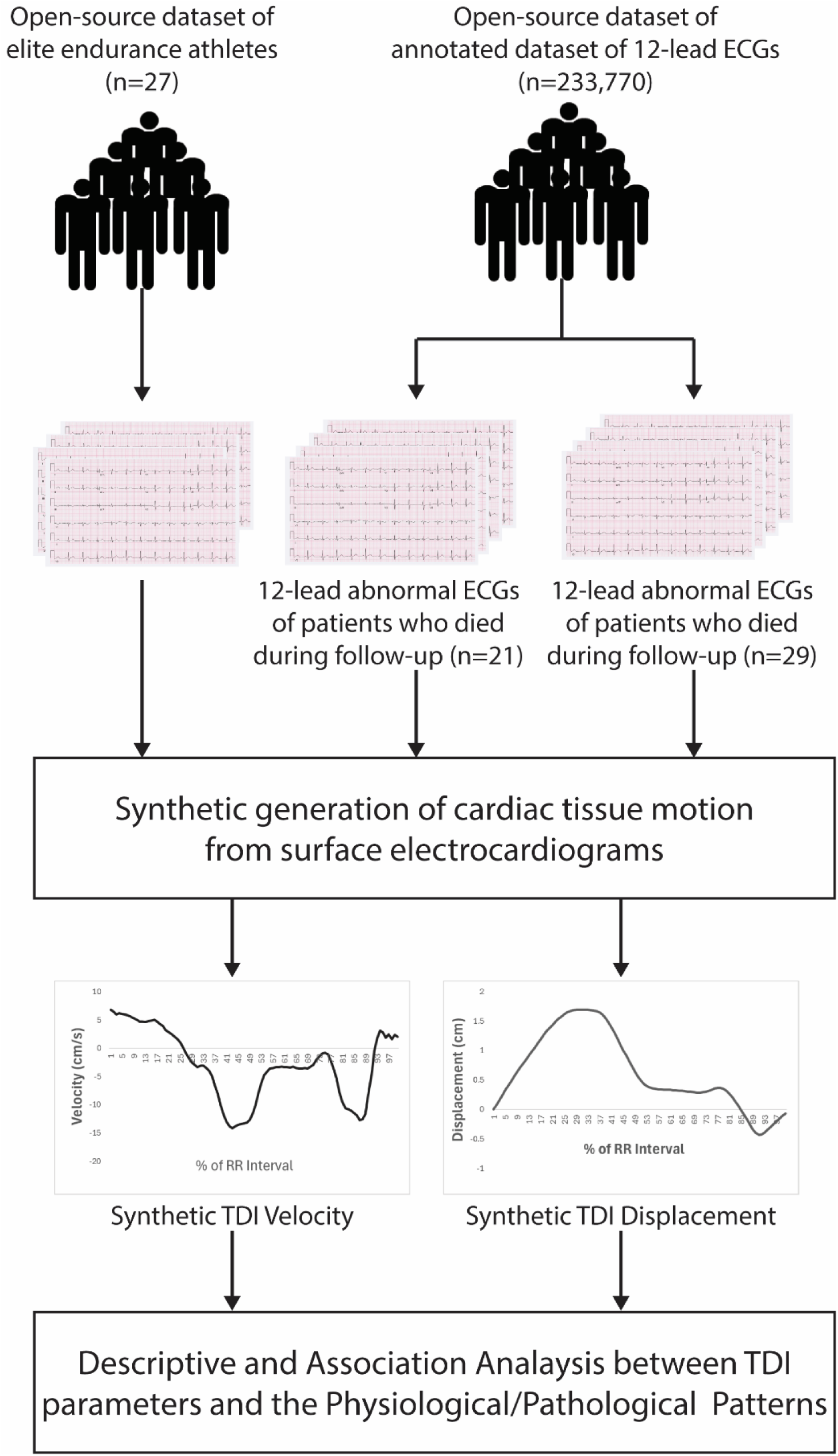
Study Design: Differentiating Athlete Adaptations from Pathological Patterns in Young Individuals. Schematic representation of the study workflow illustrating participant selection, classification into healthy athletes or individuals with pathological findings, acquisition of ECG-derived synthetic tissue Doppler imaging (TDI) waveforms, integration to obtain myocardial displacement, and statistical analysis to determine associations with physiological adaptation or mortality-linked pathological patterns in young individuals.

The participants in the athlete’s group were obtained from the PhysioNet database^16^, with recruitment conducted in accordance with ethical approvals from the University of Oslo and the Norwegian Centre for Research Data. Each athlete underwent a standard 10-second, 12-lead ECG interpreted by a sports cardiologist using international athlete-specific ECG criteria. All protocols involving athlete participants were approved by the Norwegian ethics board, with informed consent obtained.

The remaining two groups were drawn from a publicly available, annotated dataset of 12-lead ECGs (CODE-15%), which is a stratified subset of the original CODE database comprising 345,779 recordings from 233,770 patients collected by the Telehealth Network of Minas Gerais, Brazil, between 2010 and 2016 ^17^. ECGs were sampled at 400 Hz, zero-padded to 10 seconds (4096 samples) when necessary, and stored in HDF5 format with linked demographic, diagnostic, and mortality follow-up data.

The individuals with abnormal ECGs (i.e., high-risk group) were those with abnormal resting ECGs who died within two years of recording, with death presumed to be of cardiac origin. Inclusion criteria included adequate ECG quality, documented mortality during follow-up, and evidence of either right or left bundle branch block.

The individuals with normal ECGs (i.e., control group) comprised individuals with normal resting ECGs and no adverse cardiac outcomes during at least five years of follow-up. Together, these three cohorts provided distinct phenotypic profiles for assessing the discriminatory performance of synthetic TDI parameters. All participants were sourced from publicly available, open-access ECG repositories.

### ECG and Tissue Doppler Data Preprocessing

To generate synthetic TDI waveforms, raw ECG and TDI data underwent a multistep preprocessing pipeline. Original TDI images in DICOM format were converted to JPG, vectorized using automated landmark detection, and split into single RR-interval cardiac cycles. Each cycle was normalized in length, smoothed with a moving average filter, and rescaled. Corresponding 12-lead ECGs were segmented into RR intervals and resized to 512 samples, with 554 numerical ECG features (including intervals, wave morphologies, axes, amplitudes, durations, dispersion measures, and corrected QT values) extracted and standardized. Data augmentation increased training diversity by generating ten variants per sample through random cycle selection, synchronized phase shifts, segment masking, and lead removal, ensuring alignment between ECG and TDI inputs. The detailed preprocessing data is provided in Radhakrishnanet al. (2025)^1^.

### Synthetic TDI Generation and Feature Extraction

As described in our previous study^1^, we employed a pair of conditional generative adversarial networks (GANs), inspired by the pix2pix framework, to generate synthetic TDI waveforms from 12-lead ECG recordings, with each network tailored to produce septal and lateral views. We then measured the three peak velocities—s′, e′, and a′—from the ECG-derived TDI waveforms, which were subsequently used in the present study to differentiate high risk group from the athletes and the controls.

The GAN architecture consisted of a generator that produced tissue TDI waveforms from 12-lead ECGs and a discriminator that distinguished real from synthetic TDIs, with an auxiliary regressor providing additional feedback on key velocity peaks. Training incorporated adversarial, reconstruction, and auxiliary losses to preserve waveform morphology and patient-specific electromechanical features. This approach enabled the model to learn complex mappings between electrical and mechanical cardiac activity, producing synthetic TDI waveforms that closely matched real measurements in both morphology and clinical associations. In prior validation, this model showed a strong association with reference TDI measurements and demonstrated excellent ability to distinguish diastolic and systolic cardiac dysfunction^1^.

### ECG-derived Synthetic Myocardial Displacement

From the synthetic TDI velocity waveforms, myocardial displacement (mm) was computed by time-integrating the velocity signal over the cardiac cycle to estimate longitudinal myocardial motion. For each individual, lateral and septal velocity traces, along with the RR interval (in seconds), were identified. Where available, baseline drift was corrected by subtracting the row-wise mean velocity to minimize offset errors. Displacement curves were then calculated by integrating velocity over time, with the time step derived from the RR interval divided into 100 equally spaced samples representing one normalized cardiac cycle. Lateral and septal displacements were computed separately, and when both views were available, an average displacement trace was generated. For each patient, the maximum displacement values from the lateral, septal, and averaged traces were recorded. All displacement time series and maximum displacement metrics were compiled for downstream statistical analysis.

### Statistical Analysis

All statistical analyses were performed using RStudio (version 2024.03.1). Continuous variables were summarized as means ± standard deviations, while categorical variables were presented as frequencies and percentages. The Shapiro–Wilk test was used to assess the normality of continuous data distributions. Group comparisons of synthetic TDI parameters (s′, e′, a′) across the three study groups were conducted using ANOVA when normality assumptions were satisfied, and the Kruskal–Wallis test when they were violated. Post-hoc pairwise comparisons were performed using Dunn’s test with Bonferroni-adjusted p-values to control multiple comparisons. To identify TDI-based predictors distinguishing athletes from high-risk individuals with abnormal ECGs, univariate and multivariate logistic regression analyses were conducted. Prior multivariate logistic regression, we assessed collinearity among the s′, e′, a′, e′/a′ ratio, and average myocardial displacement. Pearson correlation matrices and Variance Inflation Factor (VIF) analyses are used to check for collinearity between the variables of interest. Model performance was assessed using deviance and the area under the receiver operating characteristic (ROC) curve (AUC). A two-sided p-value < 0.05 was considered indicative of statistical significance. Given the limited sample size, all findings were interpreted as exploratory and hypothesis-generating rather than confirmatory in nature.

## Results

The demographic, clinical, and cardiac functional characteristics of the study participants, stratified by group, are summarized in **Table 1**. A total of 77 individuals were categorized into three groups: high-risk group with abnormal ECGs (n = 21), controls with normal ECGs (n = 29), and athletes (n = 27). The high-risk group had the highest mean age (33.5 ± 6.1 years; range, 17–40), followed by the control group (31.1 ± 7.4 years; range, 18–43) and the athlete group (25.0 ± 4.7 years; range, 22–43). Males predominated in the high-risk group with abnormal ECGs and subsequent death (85.7%) and in the athlete group (70.4%), compared with 37.9% in the control group.

**Table 1.** Demographic, Clinical, and Cardiac Functional Characteristics of Study Participants by Group.

| Variable | High-Risk | Controls | Athletes | p-value |
| --- | --- | --- | --- | --- |
| Age (years) | 33.48 ± 6.10 | 31.07 ± 7.40 | 25.00 ± 4.70 | 0.207 |
| Males (%) | 18 (85.7) | 11 (37.9) | 19 (70.4) | 0.0015 |
| RR Interval (s) | 0.81 ± 0.16 | 0.82 ± 0.13 | 1.08 ± 0.17<sup>*#</sup> | <0.0001 |
| Lateral s' (cm/s) | 4.92 ± 2.30 | 6.00 ± 1.64 | 5.71 ± 1.42 | 0.0877 |
| Lateral e' (cm/s) | 7.30 ± 3.80 | 10.36 ± 6.04 | 7.45 ± 1.87 | 0.0906 |
| Lateral a' (cm/s) | 5.80 ± 3.50 | 4.96 ± 2.82 | 3.58 ± 2.08<sup>*#</sup> | 0.0077 |
| Lateral e'/a' ratio (cm/s) | 1.58 ± 1.04 | 2.12 ± 0.92 | 2.61 ± 1.24<sup>*</sup> | 0.0064 |
| Septal s' (cm/s) | 6.23 ± 5.38 | 6.05 ± 1.34<sup>*</sup> | 5.88 ± 0.67 | 0.0017 |
| Septal e' (cm/s) | 4.80 ± 2.28 | 6.47 ± 1.46<sup>*</sup> | 7.18 ± 1.11<sup>*</sup> | <0.0001 |
| Septal a' (cm/s) | 5.26 ± 1.71 | 6.72 ± 1.24<sup>*</sup> | 6.15 ± 0.92 | 0.0208 |
| Septal e'/a' ratio (cm/s) | 1.20 ± 1.35 | 0.99 ± 0.27 | 1.18 ± 0.17<sup>*#</sup> | 0.0133 |
| Average s' (cm/s) | 5.57 ± 3.07 | 6.02 ± 1.36<sup>*</sup> | 5.80 ± 0.79 | 0.0055 |
| Average e' (cm/s) | 6.05 ± 2.45 | 8.30 ± 3.49<sup>*</sup> | 7.32 ± 1.13<sup>*</sup> | 0.0115 |
| Average a' (cm/s) | 5.53 ± 2.09 | 5.84 ± 1.80 | 4.86 ± 1.18 | 0.087 |
| Average e'/a' ratio (cm/s) | 1.20 ± 0.57 | 1.42 ± 0.44 | 1.58 ± 0.44<sup>*</sup> | 0.016 |
| Lateral Displacement (cm) | 0.99 ± 0.61 | 1.42 ± 0.71<sup>*</sup> | 1.29 ± 0.38 | 0.0355 |
| Septal Displacement (cm) | 0.93 ± 0.40 | 1.32 ± 0.46<sup>*</sup> | 1.54 ± 0.34<sup>*</sup> | <0.0001 |
| Average Displacement (cm) | 0.83 ± 0.48 | 1.33 ± 0.55<sup>*</sup> | 1.37 ± 0.30<sup>*</sup> | 0.0016 |

\*p < 0.05 vs. high-risk, #p < 0.05 vs. controls. Values are presented as means ± standard deviations or n (%). Kruskal–Wallis with Dunn's post hoc test was used for continuous variables, RR Interval = cardiac cycle duration, s' = peak systolic velocity (cm/s), e' = early diastolic velocity (cm/s), a' = late diastolic velocity (cm/s), e'/a' ratio = ratio of early to late diastolic velocities (cm/s), Displacement = maximum myocardial displacement

In the high-risk group, right bundle branch block (RBBB) was the most prevalent finding (90.5%), followed by first-degree atrioventricular (AV) block and left bundle branch block (each 9.5%), and atrial fibrillation (4.8%). Among elite endurance athletes, abnormal ECG findings were observed in 15 participants (51.9%). The most common axis deviations were the right axis deviation in 5 athletes (18.5%) and left axis deviation in 1 (3.7%). ST-segment changes included ST elevation in 4 athletes (14.8%) and ST-Elevation Myocardial Infarction-like patterns in 4 (14.8%). Conduction abnormalities comprised AV block in 3 athletes (11.1%), right ventricular conduction delay in 2 (7.4%), nonspecific conduction abnormalities in 2 (7.4%), and incomplete RBBB in 2 (7.4%). Less frequent findings, each occurring in a single athlete (3.7%), included possible left ventricular hypertrophy, right ventricular hypertrophy, and ECG patterns suggestive of myocardial injury or infarction (possible lateral infarction, inferior infarct, lateral injury pattern, anterior infarction, anterolateral injury pattern, or pulmonary disease pattern).

The athletes had longer RR intervals compared to individuals with high-risk ECGs (p<0.001) and controls(p<0.001), corresponding to lower heart rates than both comparison groups (both p<0.001). No significant differences in RR interval or heart rate were observed between the high-risk and control groups.

In the lateral wall, a′ was lower in athletes as compared to both the high-risk and control groups (p<0.05), while the e′/a′ ratio was higher in athletes compared with the abnormal ECG with death group (p < 0.01). No significant differences were observed between groups for peak systolic velocity (s′) or peak early diastolic velocity (e′).

In the septal wall, athletes and controls demonstrated higher e′ than the high-risk group (p < 0.001). The e′/a′ ratio was higher in athletes compared with both high-risk (p < 0.05) and controls groups (p < 0.05). Peak systolic velocity (s′) c than in controls (p < 0.001)

**Figure 1** shows the comparison of synthetic TDI parameters among athletes (green), individuals in high-risk group(red), and controls (blue). On average, s′ and e′ were lower in high-risk individuals than in controls (*p* < 0.01). Athletes had a higher average e′/a′ ratio than high-risk individuals (*p* < 0.05) (Table 1, Figure 2). Furthermore, the average myocardial displacement also significantly higher in athletes as compared to high-risk individuals (p < 0.01).

**Figure 1.**
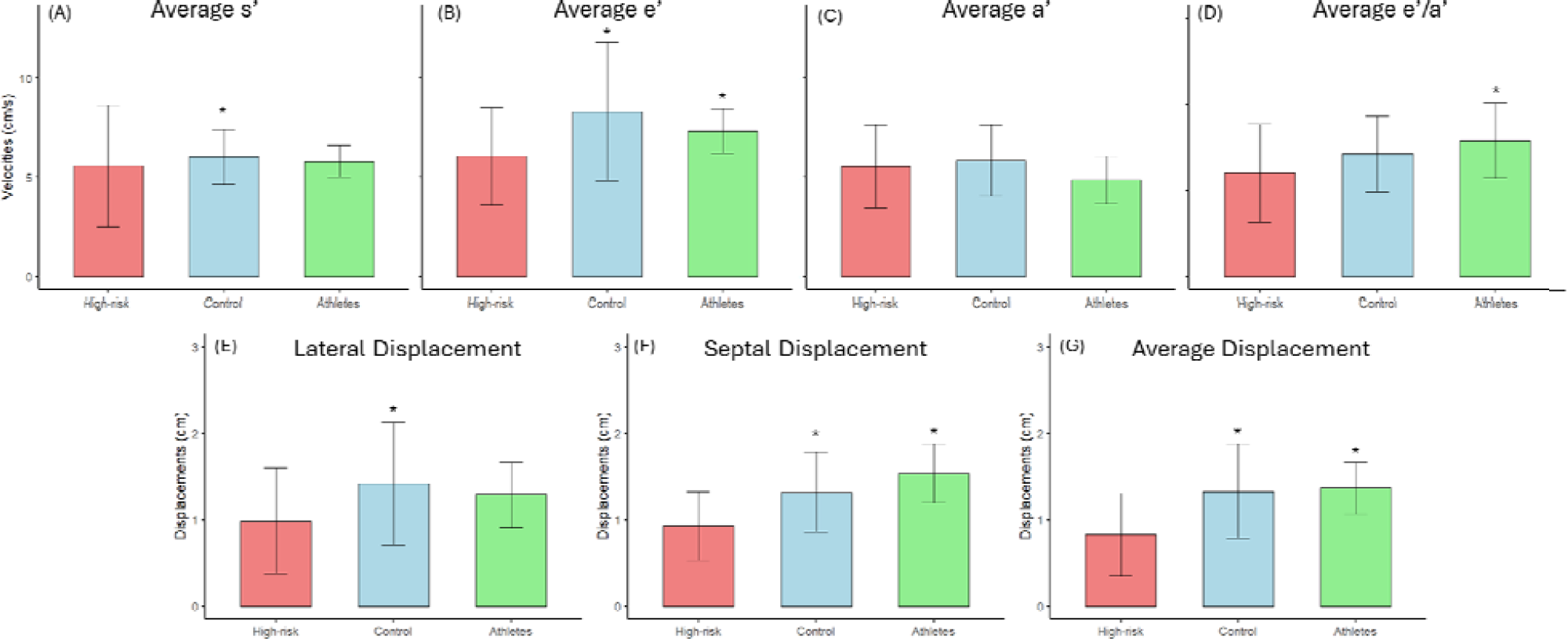
Group-wise comparison of synthetic TDI and displacement parameters. Bar plots showing (A) average peak systolic velocity (s′), (B) early diastolic velocity (e′), (C) late diastolic velocity (a′), and (D) e′/a′ ratio, as well as maximum displacement for the (F) lateral, (G) septal, and (H) averaged walls, across high-risk, controls, and athletes groups. Error bars indicate standard deviation. Statistical significance is denoted by *: p < 0.05 vs. high-risk group.

**Figure 2.**
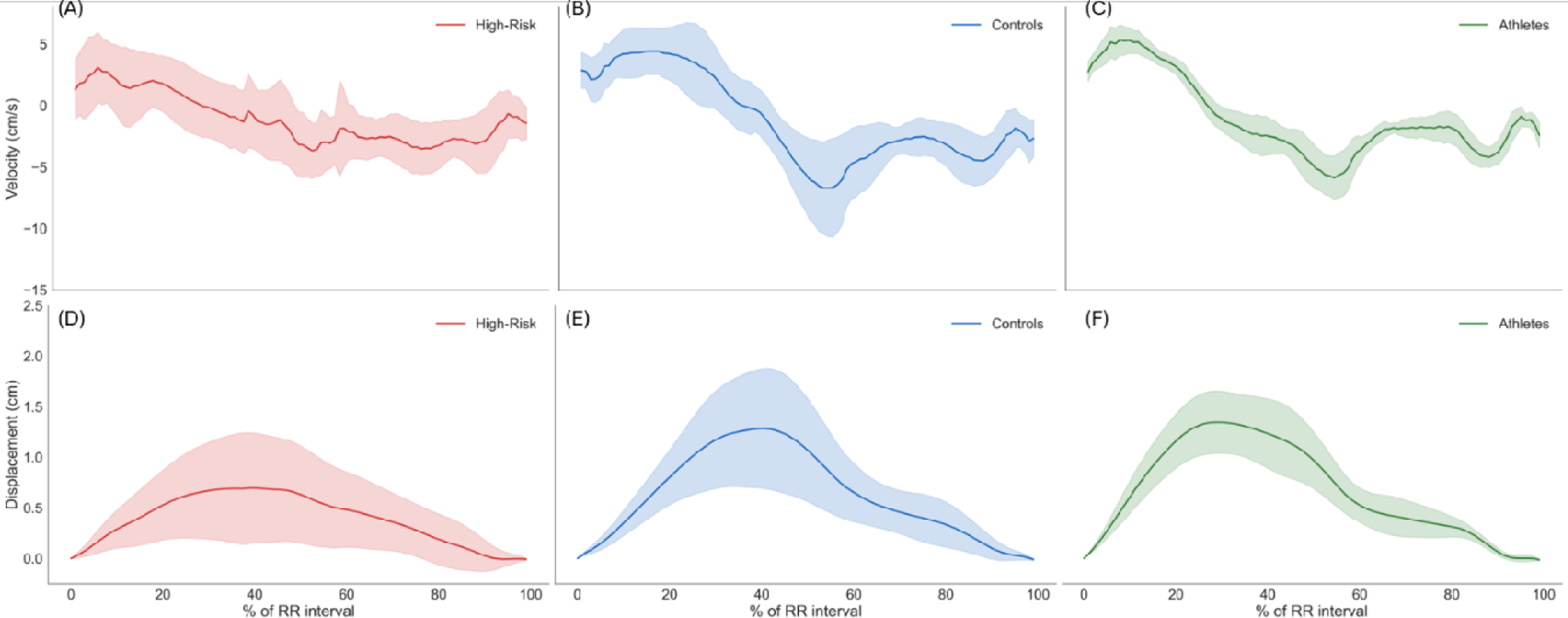
Group-wise synthetic TDI velocity and displacement waveforms. ECG-derived synthetic TDI velocity and myocardial displacement waveforms averaged across all participants in the high-risk, controls, and athletes groups. Shaded regions indicate the standard deviation, representing the variability of measurements at each time point in the cardiac cycle for both velocity and displacement waveforms.

**Figure 2** presents the average displacement curves for the three groups. For myocardial displacement, both athletes and controls showed significantly higher values than the high-risk group (p < 0.01). In the septal wall, athletes had greater displacement than the high-risk group (p < 0.001), and controls also exceeded the high-risk group (p < 0.05). In the lateral wall, controls demonstrated higher displacement than the high-risk group (p < 0.05) (**Table 1, Figure 2**). **Table 2** shows the results of univariate and multivariate logistic regression analyses for synthetic TDI parameters and their ability to distinguish athletes’ physiological patterns from pathological cardiac remodeling associated with high-risk individuals with abnormal ECGs. In univariate analysis, average early diastolic velocity (e′) was positively associated with athletes’ status (OR = 1.51, 95% CI: 1.04 – 2.20, p < 0.05; AUC = 0.750). The average e′/a′ ratio was also a strong discriminator (OR = 5.53, 95% CI: 1.32 – 23.14, p < 0.05; AUC = 0.74), while average maximum displacement demonstrated the highest discriminatory power (OR = 28.90, 95% CI: 4.24 – 197.18, p < 0.001; AUC = 0.78). No significant associations were observed for average s′ (OR = 1.06, 95% CI = 0.79 – 1.42, p = 0.71) or a′ (OR = 0.77, 95% CI = 0.54 – 1.12, p = 0.17).

**Table 2.** Univariate and multivariate logistic regression analyses comparing elite endurance athletes with individuals who had abnormal ECGs and subsequently died during follow-up.

| Variable | Univariate Analysis |  | Multivariate Analysis |  |
| --- | --- | --- | --- | --- |
|  | OR (95% CI) | p-value | OR (95% CI) | p-value |
| Average s' (cm/s) | 1.06 (0.79 – 1.42) | 0.71 | — | — |
| Average e' (cm/s) | 1.51 (1.04 – 2.20) | 0.03 | 1.15 (0.76 – 1.75) | 0.51 |
| Average a' (cm/s) | 0.77 (0.54 – 1.12) | 0.17 | — | — |
| Average e'/a' ratio (cm/s) | 5.53 (1.32 – 23.14) | 0.02 | — | — |
| Average Displacement (cm) | 28.90 (4.24 – 197.18) | <0.0001 | 24.66 (3.23 – 188.25) | 0.002 |

s' = average peak systolic velocity (cm/s), e' = early diastolic velocity (cm/s), a' = late diastolic velocity (cm/s), e'/a' ratio = ratio of average early to late diastolic velocities (cm/s), Displacement = maximum average myocardial displacement (cm) across septal and lateral annular sites.

In the multivariate analysis, collinearity assessment using Pearson correlation and variance inflation factor (VIF) revealed moderate-to-high VIF values for e′ (6.58), a′ (6.77), and the e′/a′ ratio (8.24), indicating potential multicollinearity. In contrast, s′ (1.09) and average maximum displacement (1.39) showed low VIFs, suggesting minimal collinearity. Accordingly, the final multivariate model combining e′ and maximum displacement demonstrated good discriminatory ability to distinguish athletes from high-risk individuals (OR = 24.66 (95% CI: 3.23 – 188.25), p = 0.02, AUC = 0.80), with maximum displacement alone showing an independent association (OR = 24.66, 95% CI: 3.23–188.25, p = 0.002).

## Discussion

This study demonstrates that ECG-derived synthetic TDI features can capture physiologically meaningful differences between elite endurance athletes, young individuals with abnormal ECGs who later died, and healthy controls with normal ECGs. As expected, athletes displayed higher e′ velocities, e′/a′ ratios, and septal displacement—patterns consistent with enhanced diastolic compliance and longitudinal excursion arising from chronic volume loading and augmented preload ^18,19^. In contrast, high-risk individuals exhibited reduced diastolic velocities and myocardial displacement, consistent with impaired relaxation and early systolic dysfunction ^20,21^.

Importantly, these functional patterns were derived entirely from ECG using a deep learning model, offering a physiologically grounded interpretation of electrical signals. This provides more than just classification—it adds an explainability layer to AI-based ECG interpretation. Rather than outputting a “high-risk” label without context, the model generates velocity and displacement metrics that align with established myocardial physiology. This interpretability is particularly valuable in sports cardiology, where benign ECG patterns can mimic disease, and in community screening, where imaging resources may be limited.

Our findings are consistent with prior echocardiographic TDI studies that distinguish physiological from pathological remodeling ^22,23^ yet achieve comparable discriminative performance (AUC up to 0.80) without the need for echocardiography. This suggests that synthetic TDI could serve as a cost-effective and scalable screening tool while maintaining transparency in its decision-making process. If validated in larger, diverse cohorts with paired imaging, synthetic TDI could help bridge the gap between AI detection and clinician trust by linking ECG features to interpretable functional outcomes.

Several limitations should be considered when interpreting these findings. First, the sample size was small, which limited statistical power and potentially underrepresented the full spectrum of athletic and pathological phenotypes. Second, while synthetic TDI features were previously validated against echocardiographic measurements in a separate dataset, this study did not include contemporaneous imaging for direct confirmation, meaning that the functional outputs, although physiologically plausible, remain model-derived in this cohort. Third, the data were sourced from geographically and demographically distinct populations, introducing potential variability in genetic background, training regimens, and comorbidities that may influence myocardial remodeling patterns. Fourth, the cross-sectional design precludes causal inference between ECG-derived functional differences and adverse outcomes. Finally, as with all secondary data analyses, variability in acquisition protocols and selection criteria may introduce unmeasured biases.

These limitations reinforce that our results should be interpreted as preliminary and hypothesis-generating. Future research should focus on prospective, multi-center validation with paired echocardiographic TDI, inclusion of broader age and ethnic groups, and longitudinal follow-up to confirm predictive value. Such studies will be essential to establish synthetic TDI not only as a diagnostic aid but also as a reliable and interpretable component of explainable AI pipelines for cardiac risk assessment.

## Conclusion

This pilot study demonstrates that synthetic TDI features derived from standard ECGs can provide physiologically interpretable insights into myocardial function, distinguishing benign athletic remodeling from abnormal patterns associated with increased mortality risk in young individuals. By translating ECG waveforms into myocardial velocity and displacement metrics, this approach adds an explainability layer to AI-based ECG interpretation—linking electrical findings to functional cardiac mechanics. These results suggest that scalable, imaging-free functional assessment may be feasible in both athletic and general populations, supporting early risk stratification where imaging is unavailable. Prospective validation with paired echocardiography and larger, diverse cohorts will be critical to confirm diagnostic accuracy, generalizability, and clinical utility.

## Data Availability

All data produced in the present work are contained in the manuscript

## References

1. Radhakrishnan A, Yanamala N, Jamthikar A, et al. Synthetic generation of cardiac tissue motion from surface electrocardiograms. Nature Cardiovascular Research. 2025;4(4):445–457.

2. Sengupta S, Jain R, Burkule N, Olet S, Khandheria BK. Myocardial work index: A novel method for assessment of myocardial function in south asian recreational athletes. Journal of Patient-Centered Research and Reviews. 2020;7(2):147.

3. Wagner E. 10 Heart Disease Statistics Every Clinician Can Use to Help Their Patients. Heart Disease. 2024.

4. Di Cesare M, Perel P, Taylor S, et al. The heart of the world. Global heart. 2024;19(1):11.

5. Vähätalo J, Holmström L, Pakanen L, et al. Coronary artery disease as the cause of sudden cardiac death among victims< 50 years of age. The American Journal of Cardiology. 2021;147:33–38.

6. Harmon KG, Asif IM, Klossner D, Drezner JA. Incidence of sudden cardiac death in National Collegiate Athletic Association athletes. Circulation. 2011;123(15):1594–1600.

7. Hayashi M, Shimizu W, Albert CM. The spectrum of epidemiology underlying sudden cardiac death. Circulation research. 2015;116(12):1887–1906.

8. Maron BJ, Doerer JJ, Haas TS, Tierney DM, Mueller FO. Sudden deaths in young competitive athletes: analysis of 1866 deaths in the United States, 1980–2006. Circulation. 2009;119(8):1085-1092.

9. Update A. Heart disease and stroke statistics–2017 update. Circulation. 2017;135(e146-e603):1.

10. Barakat MF, Chehab O, Kaura A, et al. Tissue doppler-derived left ventricular systolic velocity is associated with lethal arrhythmias in cardiac device recipients irrespective of left ventricular ejection fraction. Journal of the American Society of Echocardiography. 2020;33(12):1509–1516.

11. Sengupta PP, Mohan JC, Pandian NG. Tissue Doppler echocardiography: principles and applications. Indian heart journal. 2002;54(4):368–378.

12. Yu C-M, Sanderson JE, Marwick TH, Oh JK. Tissue Doppler imaging: a new prognosticator for cardiovascular diseases. Journal of the American College of Cardiology. 2007;49(19):1903–1914.

13. Santoro A, Alvino F, Antonelli G, et al. Endurance and strength athlete’s heart: analysis of myocardial deformation by speckle tracking echocardiography. Journal of cardiovascular ultrasound. 2014;22(4):196.

14. Ho CY, Sweitzer NK, McDonough B, et al. Assessment of diastolic function with Doppler tissue imaging to predict genotype in preclinical hypertrophic cardiomyopathy. Circulation. 2002;105(25):2992–2997.

15. Nagueh SF. Tissue Doppler imaging for the assessment of left ventricular diastolic function. J Cardiovasc Ultrasound. 2008;16(3):76–79.

16. Goldberger AL, Amaral LA, Glass L, et al. PhysioBank, PhysioToolkit, and PhysioNet: components of a new research resource for complex physiologic signals. circulation. 2000;101(23):e215–e220.

17. Ribeiro AH, Paixao G, Lima EM, et al. CODE-15%: A large scale annotated dataset of 12-lead ECGs. Zenodo, Jun. 2021;9:10–5281.

18. Lovic D, Narayan P, Pittaras A, Faselis C, Doumas M, Kokkinos P. Left ventricular hypertrophy in athletes and hypertensive patients. The Journal of Clinical Hypertension. 2017;19(4):413–417.

19. D’Andrea A, Cocchia R, Riegler L, et al. Left ventricular myocardial velocities and deformation indexes in top-level athletes. Journal of the American Society of Echocardiography. 2010;23(12):1281–1288.

20. Kadappu KK, Thomas L. Tissue Doppler imaging in echocardiography: value and limitations. *Heart*, Lung and Circulation. 2015;24(3):224–233.

21. Møller JE, Egstrup K, Køber L, Poulsen SH, Nyvad O, Torp-Pedersen C. Prognostic importance of systolic and diastolic function after acute myocardial infarction. American heart journal. 2003;145(1):147–153.

22. Caselli S, Maron MS, Urbano-Moral JA, Pandian NG, Maron BJ, Pelliccia A. Differentiating left ventricular hypertrophy in athletes from that in patients with hypertrophic cardiomyopathy. The American journal of cardiology. 2014;114(9):1383–1389.

23. Zaidi A, Sheikh N, Jongman JK, et al. Clinical differentiation between physiological remodeling and arrhythmogenic right ventricular cardiomyopathy in athletes with marked electrocardiographic repolarization anomalies. Journal of the American College of Cardiology. 2015;65(25):2702–2711.

